# When it is available, will we take it? Public perception of hypothetical COVID-19 vaccine in Nigeria

**DOI:** 10.1101/2020.09.24.20200436

**Authors:** Yusuff Adebayo Adebisi, Aishat Jumoke Alaran, Obasanjo Afolabi Bolarinwa, Wuraola Akande-Sholabi, Don Eliseo Lucero-Prisno

**Affiliations:** Department of Clinical Pharmacy and Pharmacy Administration, Faculty of Pharmacy, University of Ibadan, Ibadan, Nigeria; Faculty of Pharmaceutical Sciences, University of Ilorin, Ilorin, Nigeria; Department of Public Health Medicine, School of Public Health and Nursing, University of KwaZulu-Natal, Durban, South Africa; Department of Global Health and Development, London School of Hygiene and Tropical Medicine, London, United Kingdom

**Keywords:** COVID-19, Vaccine Acceptance, COVID-19 Vaccine, Vaccine Hesitancy, Nigeria

## Abstract

**Introduction:** COVID-19 pandemic is a global public health threat facing mankind. There is no specific antiviral treatment for COVID-19, and no vaccine is currently available. This study aimed to understand the perception of the public towards a hypothetical COVID-19 vaccine in Nigeria.

**Method:** We conducted a cross-sectional survey in August 2020 across the 36 states of Nigeria using an online questionnaire. The questionnaire includes sections on the demographic characteristics of the respondents and their perception regarding a hypothetical COVID-19 vaccine. A total of 517 respondents completed and returned the informed consent along with the questionnaire electronically. Data were coded and abstracted into the Microsoft Excel spreadsheet and loaded into the STATA 14 software for final analysis.

**Results:** The results showed that more than half of the respondents were male 294 (56.9%). Most of the respondents (385, 74.5%) intend to take the COVID-19 vaccine when it becomes available. Among the 132 respondents that would not take the COVID-19 vaccine, the major reason for non-acceptance was unreliability of the clinical trials 49 (37.1%), followed by the belief that their immune system was sufficient to combat the virus 36 (27.3%). There were significant association between the age of the respondents and the COVID-19 vaccine acceptance *(P-value=0*.*00)* as well as geographical location and COVID-19 vaccine acceptance *(P-value=0*.*02)*.

**Conclusion:** It was observed that most of the respondents were willing to take the COVID-19 vaccine. Our findings reiterate the need to reassure the public that any vaccine that becomes available will be safe and effective. In addition, there is a need for the national health authorities to ensure the public trust is earned and all communities, including the marginalized populations, are engaged properly to ensure an optimal COVID-19 vaccine acceptance.

## INTRODUCTION

The novel coronavirus disease 2019 (COVID-19) caused by the Severe Acute Respiratory coronavirus 2 was first discovered in Wuhan Animal Market, Hubei Province, China in December 2019 [1]. Since its discovery, it has spread to more than 200 countries around the world and has been declared a pandemic by the World Health Organization [2]. COVID-19 has had unprecedented impacts on the health and well-being of people all over the world, as well as the economy of many countries [3, 4]. In Nigeria, the index case was reported on 27 February 2020, and more than 50,000 cases and over a thousand deaths have been documented since then [5].

As with past outbreaks, the novel nature of the current coronavirus outbreak implied that no definitive therapy existed for its treatment, instead, empirical therapies are being employed to manage the disease [1]. The rapid spread of the virus and continuous increasing number of cases alongside the partial and/or total lockdown protocols in most countries necessitate the urgent development of accurate diagnostic methods, effective treatments, and vaccines for the disease [6]. The long-term solution to COVID-19, however, would most likely be a safe, globally implemented vaccination program with a broad range of clinical and socioeconomic benefits [7]. It is of point to note that vaccination is one of the greatest achievements of modern medicine and it is the greatest human intervention besides clean water and sanitation [8].

Major infections such as smallpox and rinderpest have been eradicated worldwide due to vaccines, while polio has almost been eradicated with the exception of Afghanistan and Pakistan where it is still endemic [9,10,11]. The impact of vaccination on vaccine-preventable diseases cannot be overemphasized. The incidence and prevalence of diseases such as cervical cancer, hepatitis, yellow fever, tuberculosis, cholera, and tetanus, among others, have been severely reduced due to vaccine availability [12]. Vaccination plays such important roles in the fight against disease eradication and elimination, control of mortality, morbidity and complications, mitigation of disease severity, prevention of infection and even protection of unvaccinated population through herd immunity [8]. Thus, the availability of COVID-19 vaccine(s) will drastically change the course of the pandemic. As of 9 September 2020, 35 vaccine candidates are in clinical evaluation and 145 vaccine candidates are in preclinical evaluation [13].

Despite the immense benefits that vaccination has offered since the first discovery of the smallpox vaccine by Edward Jenner till date, saving millions of lives globally and doing so at a comparatively low cost, vaccine hesitancy has always plagued this great discovery. Vaccine hesitancy is “the reluctance or refusal to vaccinate despite the availability of vaccines” [14]. It was classified as one of the top ten threats to global health by the World Health Organization in 2019 [14]. Vaccine hesitancy is a complex phenomenon, with a growing continuum between vaccine acceptance and refusal. Despite the proven effectiveness and safety of vaccines, an increasing number of individuals perceive vaccines as unsafe and unnecessary [15]. In recent times, there has been a steady decline in vaccine coverage and an increase in the occurrence of vaccine-preventable diseases. For instance, there has been a 30% rise in measles cases globally. Vaccine hesitancy is believed to contribute greatly to this [14,15].

Nigeria is the most populated country in Africa and has a convoluted history of vaccine hesitancy. Vaccination coverage in Nigeria has continuously dropped since its peak of 81.5% in the 1990s, and by 2013, only 25% of children under the age of 2 were fully vaccinated [16] The 2003/2004 polio vaccine refusal in Nigeria had a far-reaching effect. It increased the incidence of polio by many folds in Nigeria and contributed to outbreaks of polio across three other continents [17]. Vaccine hesitancy could have a direct and wide-reaching effect on the acceptance of COVID-19 vaccine(s) by individuals in the community as it confers threat not only on the hesitant individual but on the community as a whole, as delays and refusals would make it impossible for communities to reach the threshold of vaccine uptake necessary for the conferment of herd immunity. While the focus of attention currently is on developing a vaccine to protect the population against COVID-19, stakeholders should prepare for the next challenge: vaccine adoption (access and acceptance) among the public. Our study aims to understand the perception of the public in Nigeria towards the hypothetical COVID-19 vaccine.

## Methods

### Study design and sampling technique

We conducted a cross-sectional survey among males and females living in Nigeria at the time of data collection and aged 15 years and older. We used a non-probability convenient sampling technique to recruit the respondents, who were required to fill the questionnaire within the first-one-month period stipulated for the study as used in previous studies [18,19]. The inclusion criteria were being social media users and having access to an internet connection to fill out the online questionnaire. We excluded individuals who do not consent to participate in the study and younger than 15 years of age [18].

### Study instrument and administration

The researchers developed the study questionnaire in several stages of drafts and reviews and with contributions from survey research experts. We conducted a pretest among ten respondents from the six geopolitical zones in Nigeria using the drafted questionnaire. The pretest was undertaken to examine readability, comprehensibility, and face validity. The final questionnaire comprises sections on the demographic characteristics of the respondents (independent variables), the intention to accept a COVID-19 vaccine was measured using a one-item question (If a vaccine against COVID-19 becomes available, would you take it? – with a Yes or No option), another question on the reasons for not intending to take the vaccine and a question on whether they have reservations towards vaccination - with a Yes or No option (outcome variables). The final questionnaire was entered into an online survey system, and a link to the electronic questionnaire was shared with respondents across the 36 states of Nigeria using social media platforms, specifically WhatsApp and Facebook, as used in a previous survey [18]. In addition to this, we urged our social media networks to share the electronic questionnaire with their networks. This was done to facilitate the achievement of more respondents. Data collection was conducted in August 2020. A total of 517 respondents completed and returned informed consent along with the questionnaire electronically via the online survey.

### Data analysis

Data were coded and abstracted into the Microsoft Excel spreadsheet and loaded into the STATA 14 software for final analysis. Simple descriptive analysis, including frequencies and percentages, was computed for demographic characteristics, and the reasons for non-acceptance of the COVID-19 vaccine, intention to take vaccine and reservations toward vaccination were presented with bar charts. A Chi-square test was carried out to determine the significant level of association and the relationship between the independent variables and outcome variables with statistical significance defined at P < 0.05.

## Results

### Socio-demographic characteristics of the respondents

Table 1 showed that more than half of the respondents were male, 294 (56.9%). The majority 478 (92.5%) of the respondents were between 16 to 30 years of age, while most of these respondents were students 302 (58.4%). The geographical location of the respondents showed that the majority of the respondents were from South-West 298 (57.6%).

**Table 1:** Socio-demographic distribution of respondents.

| Variable | *N=517* |  |
| --- | --- | --- |
| Sex | Frequency | Percentage (%) |
| Female | 223 | 43.1 |
| Male | 294 | 56.9 |
| Age group (years) |  |  |
| 16-30 | 478 | 92.5 |
| 31-45 | 29 | 5.6 |
| 46-60 | 6 | 1.2 |
| 61+ | 4 | 0.7 |
| Employment |  |  |
| Employed | 179 | 34.6 |
| Student | 302 | 58.4 |
| Unemployed | 36 | 7.0 |
| Geo-political Zone |  |  |
| North-Central | 77 | 14.9 |
| North-East | 6 | 1.2 |
| North-West | 12 | 2.3 |
| South-South | 69 | 13.4 |
| South-East | 55 | 10.6 |
| South-West | 298 | 57.6 |
| **Education Level** |  |  |
| Graduate | 171 | 33.1 |
| Postgraduate | 52 | 10.0 |
| Secondary | 15 | 2.9 |
| Undergraduate | 279 | 54.0 |

### Percentage distribution of respondents’ reservation toward vaccination, acceptance of COVID-19 vaccine and reasons for non-acceptance of COVID-19 vaccine

The results showed that 385 (74.6%) are willing to receive COVID-19 vaccine. 124 (24.0%) have reservations toward vaccination (Figure 1). The major reason for non-acceptance of the COVID-19 vaccine among our respondents is the unreliability of the clinical trials 49 (37.1%), followed by the belief that their immune system is enough to combat the virus 36 (27.3%) as shown in Figure 2.

**Figure 1:**
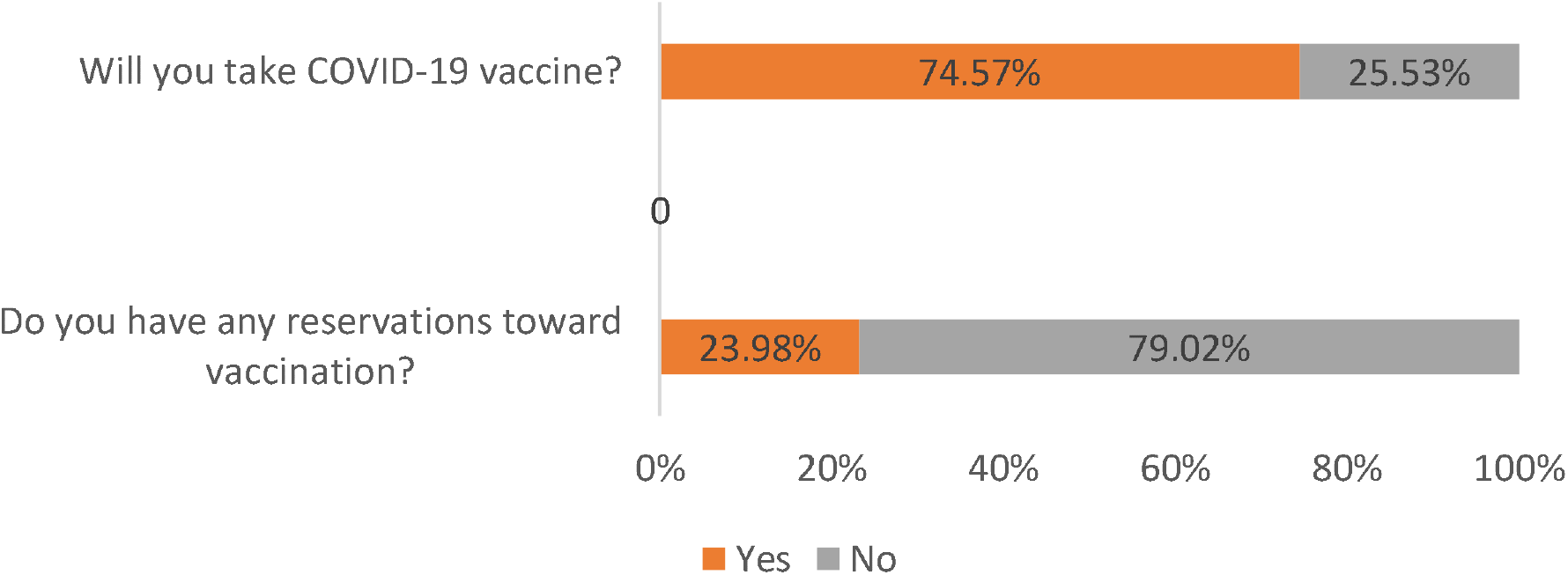
Percentage distribution of respondents reservations toward vaccination and acceptance of COVID-19 vaccine [n=517]. Percentage distribution of respondent’s reservations toward vaccination and acceptance of the COVID-19 vaccine.

**Figure 2:**
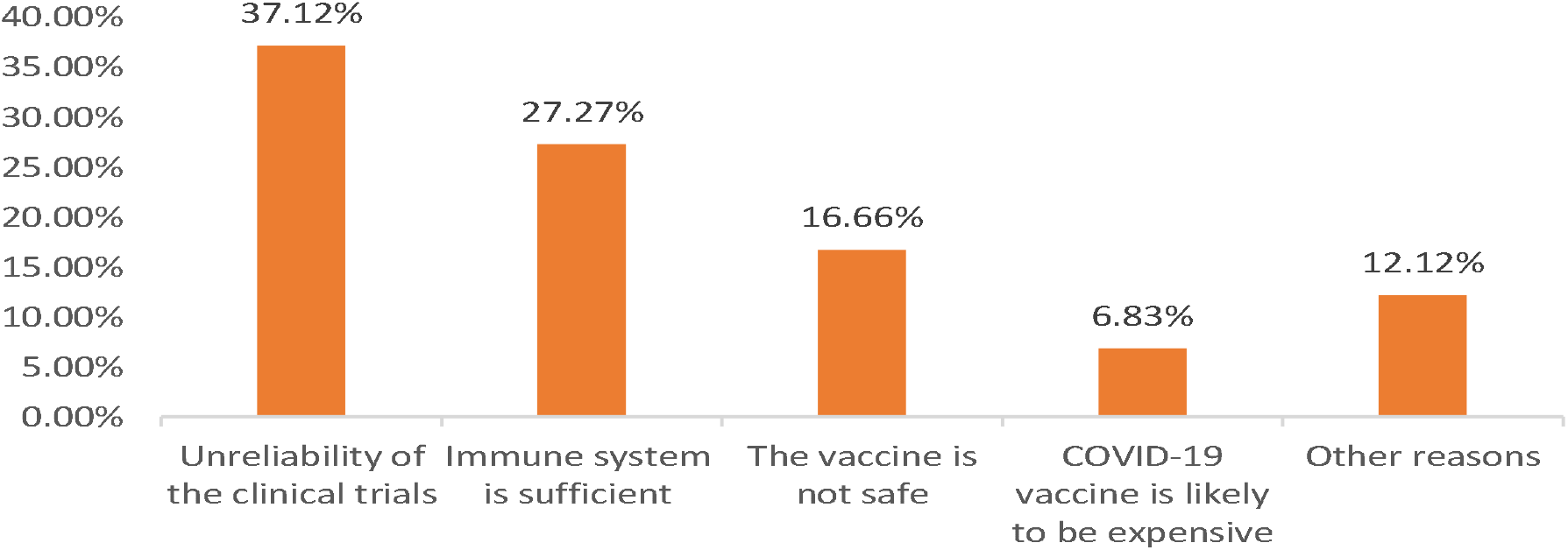
Reasons for non-acceptance of COVID-19 vaccine by our respondents [n=132]. Reasons for non-acceptance of the COVID-19 vaccine by our respondents.

### Association between selected socio-demographic variables, having reservations toward vaccination and COVID-19 vaccine acceptance

Our result revealed having reservations toward vaccination [**χ2**=0.10 *P-value=0*.*76*] and COVID-19 vaccine acceptance [**χ2**=1.53 P*-value=0*.*22*] are not statistically associated with the sex of respondents. Both COVID-19 vaccine acceptance [***χ2****=24*.*33 P-value=0*.*00*] and reservation towards vaccination [***χ2****= 19*.*04 P-value=0*.*00*] are significantly associated with the age group. Only COVID-19 vaccine acceptance [***χ2****=13*.*78 P-value=0*.*02*] is significantly associated to the respondents’ geopolitical zone. Having reservation towards vaccination [***χ2****=12*.*01 P-value=0*.*00]* is also significantly associated to the respondents’ education level (Table 2)

**Table 2:** Association between selected socio-demographic variables, vaccine reservation and COVID-19 vaccine acceptance.

| Variable N=517 | Do you have any reservations towards vaccination?<br>(Reservations toward vaccination) |  | Will you take COVID-19 vaccine? (COVID-19 vaccine acceptance) |  |
| --- | --- | --- | --- | --- |
|  | No<br>n (%) | Yes<br>n (%) | No<br>n (%) | Yes<br>n (%) |
| Sex |  |  |  |  |
| Female (223) | 171 (76.7) | 52 (23.3) | 63 (28.3) | 160 (71.7) |
| Male (294) | 222 (75.5) | 72 (24.5) | 69 (23.5) | 225 (76.5) |
| | $\chi^2=0.10$ $P\text{-value}=0.76$ | | $\chi^2=1.53$ $P\text{-value}=0.22$ | |
| **Age group (years)** |  |  |  |  |
| 16-30 (478) | 369 (77.2) | 109 (22.8) | 113 (23.6) | 365 (76.4) |
| 31-45 (29) | 22 (75.9) | 7 (24.1) | 10 (34.5) | 19 (65.5) |
| 46-60 (6) | 2 (33.3) | 4 (66.7) | 5 (83.3) | 1 (16.7) |
| 61+ (4) | 0 (0.0) | 4 (100.0) | 4 (100.0) | 0 (0.0) |
| | $\chi^2=19.04$ $P\text{-value}=0.00^*$ | | $\chi^2=24.33$ $P\text{-value}=0.00^*$ | |
| **Employment** |  |  |  |  |
| Employed (179) | 125 (69.8) | 54 (30.2) | 46 (25.7) | 133 (74.3) |
| Student (302) | 243 (80.5) | 59 (19.5) | 74 (24.5) | 228 (75.5) |
| Unemployed (36) | 25 (69.4) | 11 (30.6) | 12 (33.3) | 24 (66.7) |
| | $\chi^2=7.88$ $P\text{-value}=0.02^*$ | | $\chi^2=1.32$ $P\text{-value}=0.52$ | |
| **Geo-political** |  |  |  |  |
| North-Central (77) | 55 (71.4) | 22 (28.6) | 20 (26.0) | 57 (74.0) |
| North-East (6) | 6 (100.0) | 0 (0.0) | 2 (33.3) | 4 (66.7) |
| North-West (12) | 7 (58.3) | 5 (41.7) | 5 (41.7) | 7 (58.3) |
| South-South (69) | 51 (73.9) | 18 (26.1) | 14 (20.3) | 55 (79.7) |
| South-East (55) | 41 (74.5) | 14 (25.5) | 24 (43.6) | 31 (56.4) |
| South-West (298) | 233 (78.2) | 65 (21.8) | 67 (22.5) | 231 (77.5) |
| | $\chi^2=5.84$ $P\text{-value}=0.32$ | | $\chi^2=13.78$ $P\text{-value}=0.02^*$ | |
| **Education Level** |  |  |  |  |
| Graduate (171) | 129 (75.4) | 42 (24.6) | 39 (22.8) | 132 (77.2) |
| Postgraduate (52) | 30 (57.7) | 22 (42.3) | 20 (38.5) | 32 (61.5) |
| Secondary (15) | 11 (73.3) | 4 (26.7) | 6 (40.0) | 9 (60.0) |
| Undergraduate (279) | 223 (80.0) | 56 (20.0) | 67 (24.0) | 212 (76.0) |
| | $\chi^2=12.01$ $P\text{-value}=0.00^*$ | | $\chi^2=7.22$ $P\text{-value}=0.06$ | |
| **Total** | **393 (76.0)** | **124 (24.0)** | **132 (25.5)** | **385 (74.5)** |

\* significant at $P<0.05$

## DISCUSSION

This study is the first in Nigeria to assess the public perception towards the hypothetical COVID-19 vaccine. It has been documented that the public perception of vaccination governs the effectiveness of vaccination programs [20]. There is no specific antiviral treatment for COVID-19, and no vaccine is currently available [1]. Vaccination, one of the greatest advances in medicine, is one of the most effective tools for reducing the burden of infectious diseases. Over the last 20 years, worldwide, vaccination programs for polio, whooping cough, diphtheria, and measles have significantly reduced the prevalence of these diseases [9]. Despite the benefit vaccination reap for public health, this fundamental effort for disease control still faces major obstacles globally, and Nigeria is not an exception [21]. It has been noted that one of the major obstacles to vaccine acceptance is the public perception of the relative risks and benefits of vaccination [22].

Findings from our study revealed that there is no significant association between COVID-19 vaccine acceptance and sex. This contrasts with a multi-national survey in Europe, where there was a significant association between the two variables and a significantly higher proportion of males were willing to get vaccinated [23]. Even though there is no significant difference in the willingness to take the COVID-19 vaccine and sex, males are slightly willing to take the COVID-19 vaccine than females. Our results also revealed a significant association between age group and COVID-19 vaccine acceptance, with respondents age 16 to 30 more willing to take the COVID-19 vaccine. One might argue that the group who currently disagreed with taking the COVID-19 vaccine may be the most relevant. However, these sets of people can easily be persuaded to get vaccinated to achieve herd immunity; if they are informed of the benefits, vaccination can reap for public health.

Regarding the COVID-19 vaccine acceptance, about 74% of our respondents agreed to take the COVID-19 vaccine when it is available. This is higher than what was reported in South Africa (64%), Russia (54%), Poland (56%), Hungary (56%), and France (59%), according to a recent World Economic Forum’s Ipsos survey of nearly 20,000 adults on whether they will take COVID-19 vaccine or not when it is available [24]. However, the willingness to take the COVID-19 vaccine was higher in China (97%), Brazil (88%), Australia (88%), and India (87%) [24] compared to our study. A study in Indonesia also reported that for a 95% effective COVID-19 vaccine, 93.3% would take it [25]. Another study in Malaysia revealed that 94.3% of the respondents would take the COVID-19 vaccine [26].

Finding from our study also revealed that geographical location and acceptance of the COVID-19 vaccine are significantly associated. This is evident in that respondents from the Southern part of the country are likely to take the COVID-19 vaccine compared to the Northern part of the country. This is not surprising because previous studies have reported high levels of vaccine refusal in the northern part of Nigeria [27,28]. For instance, in 2003, five northern Nigerian states boycotted the oral polio vaccine due to fears that it was unsafe [17]. However, these findings don’t necessarily imply that refusal would be higher in Northern Nigeria; it thus strengthens the need to reassure the public of the safety of the COVID-19 vaccine irrespective of their geographical location. The acceptance level of the COVID-19 vaccine, according to our study, is lower than that of the hypothetical Ebola vaccine (80%) [29] and malaria vaccine (96%) [30] in previous studies conducted in Nigeria.

Furthermore, about 25% of our respondents disagreed with taking the COVID-19 vaccine when it is available. This is higher than what was reported in China (3%), Brazil (12%), Australia (12%), India (13%), Malaysia (15%), Great Britain (15%), Saudi Arabia (16%), South Korea (16%), Peru (21%), and Canada (24%) according to a recent World Economic Forum’s Ipsos survey of nearly 20,000 adults on whether they will take COVID-19 vaccine or not when it is available [24]. However, the unwillingness to take the COVID-19 vaccine was higher in Russia (47%), Poland (45%), Hungary (44%), France (41%), South Africa (36%), Sweden (33%), United States (33%), Germany (33%) and Italy (33%) [24] compared to our study.

Findings from our study also revealed that the top three reasons for non-acceptance of the COVID-19 vaccine are the unreliability of clinical trials, the immune system is enough to combat COVID-19, and that the COVID-19 vaccine is not safe. This is similar to what was reported by the Ipsos survey, where the top 3 reasons were “worry about side effects”, “doubt about the vaccine effectiveness”, and “perception of not being enough at risk from COVID-19” [24]. Another multi-national study in Europe revealed that more than half said they were concerned about the potential side effects of the vaccine [23]. This is not surprising in that all these findings have been found in literature as some of the reasons for vaccine hesitancy [31,32]. It has also been documented in the literature that vaccine adoption is a sum of vaccine access and acceptance [33]. Interestingly, one of the reasons for the non-acceptance of the COVID-19 vaccine is the vaccine’s perceived high cost according to our findings.

Findings from this study revealed 25% of our respondent will not accept the COVID-19 vaccine. Thus, there is a possible implication of influencing others with their perception of the COVID-19 vaccine leading to more people refusing the vaccine. This could lead to widespread refusal of COVID-19 vaccine due to eroded public trust with the negative information regarding the COVID-19 vaccine. The estimated reproductive number of COVID-19 for Nigeria has been determined to be 2.63 [34]. Going by the formula Q = 1 – 1/R, where Q is the herd immunity threshold and R is the reproductive number [35], an estimated 62% or more of the Nigerian population must be vaccinated to achieve herd immunity. For Nigeria to achieve this, national health authorities need to devise means to ensure the public trust is earned and all communities, including the marginalized populations, must be engaged properly to ensure an increase in vaccine acceptance. In addition, all information regarding the vaccine must be made public, quality control and assurance must also be the priority of the health authorities.

Access is also an important factor to be considered as regards the COVID-19 vaccine, and it is essential to translate the willingness to be vaccinated into actual vaccination decisions when the vaccine becomes available. Even though our study did not assess the willingness to pay for the COVID-19 vaccine, equitable access to vaccination is much-needed to quickly achieve herd immunity.

## LIMITATIONS

Our study is not without its limitations. Generalization of the survey results should be avoided because the pattern of questionnaire distribution may influence the outcome. We used the social media platform, so it may omit older adults, people from lower socioeconomic classes, certain geographical locations, lower educational attainment, and those who were illiterates as well as people who did not have access to the internet. Besides, response/social bias is also a possibility which is not uncommon in self-administered questionnaire research. Finally, acceptance was assessed using a potential (hypothetical) vaccine, which may differ from the respondents’ revealed preferences when the vaccine becomes available. Thus, future study may need to put all these gaps into consideration to ensure a far-reaching conclusion in this regard.

## CONCLUSION

Even though most of our respondents are willing to take the COVID-19 vaccine, our findings reiterate the need to reassure the public that any vaccine which becomes available will be safe and effective. Otherwise, there is a risk of a reversal of hard-won achievement of building public trust in Nigeria’s vaccination programme, potentially compromising reaching community immunity. Public trust is important in promoting public health and plays an essential role in the public’s compliance with vaccination programs and other health interventions. However, if the public trust is eroded, false information can spread, leading to the rejection of health interventions with a major threat to public health. Besides, national health authorities, stakeholders and policymakers in Nigeria need to ensure that access to the COVID-19 vaccine is equitable when it becomes available.

## Data Availability

Available

## Acknowledgment

We would like to thank all the respondents of the study for their participation. We also appreciate the reviewers for their insightful comments.

## Authors contribution

Yusuff Adebayo Adebisi and Aishat Jumoke Alaran conceptualized the study. Yusuff Adebayo Adebisi, Obasanjo Afolabi Bolarinwa and Aishat Jumoke Alaran wrote the first draft of the paper and performed data analysis. Wuraola Akande-Sholabi and Don Eliseo Lucero-Prisno III critically reviewed and suggested important improvements to the manuscript. All authors read and approved the final manuscrip.

## Ethics Approval

Ethical approval was not necessary because the study focussed on perception of our respondents and no confidential information was asked.

## Consent for publication

Not applicable

## References

1. Ge H, Wang X, Yuan X, Xiao G, Wang C, Deng T, et al., 2020. The epidemiology and clinical information about COVID-19. Eur J Clin Microbiol Infect Dis. 39(6). 1011–1019.

2. Cucinotta D, Vanelli M, 2020. WHO declares COVID-19 a pandemic. Acta Biomedica, 91(1), 157–160.

3. Lucero-Prisno DE, Adebisi YA, Lin X, 2020. Current efforts and challenges facing responses to 2019-nCoV in Africa. Glob Health Res Policy, 5:21.

4. Ogunkola IO, Adebisi YA, Imo UF, Odey GO, Esu E, Lucero-Prisno DE, 2020. Rural communities in Africa should not be forgotten in responses to COVID-19. Int J Health Plann Manag. 10.1002/hpm.3039. Advance online publication.

5. Nigeria Centre for Disease Control. (2020). COVID-19 NIGERIA. Retrieved September 5, 2020, from NCDC website: https://covid19.ncdc.gov.ng/report/

6. Shih HI, Wu CJ, Tu YF, Chi CY, 2020. Fighting COVID-19: A quick review of diagnoses, therapies, and vaccines. Biomed J. 2020;S2319-4170(20)30085-8.

7. Schaffer Deroo S, Pudalov NJ, Fu LY, (2020). Planning for a COVID-19 Vaccination Program. JAMA. 323(24);2458–2459.

8. Andre FE, Booy R, Bock HL, Clemens J, Datta SK, John TJ, Lee BW, Lolekha S, Peltola H, Ruff TA, Santosham M, Schmitt HJ, 2008. Vaccination greatly reduces disease, disability, death and inequity worldwide. Bull World Health Organ. https://www.who.int/bulletin/volumes/86/2/07-040089/en/ Accessed date: September 12, 2020

9. Greenwood B, 2014. The contribution of vaccination to global health: Past, present and future. Philosophical Transactions of the Royal Society B: Biological Sciences,. 369(1645).

10. Polio Global Eradication Initiative. (2020). Endemic Countries. http://polioeradication.org/where-we-work/polio-endemic-countries/ Accessed date: September 12, 2020

11. Ahmadi A, Essar MY, Lin X, Adebisi YA, Lucero-Prisno DE. 2020. Polio in Afghanistan: The Current Situation amid COVID-19. Am J Trop Med Hyg. Aug 27. doi: 10.4269/ajtmh.20-1010. Epub ahead of print.

12. Miller MA, Sentz JT, 2006. Vaccine-preventable diseases. In Jamison DT, Feachem RG, Makgoba MW, et al. (Eds.), Disease and mortality in sub Saharan Africa. 2nd ed. Washington (DC): World Bank. Chapter 12.

13. World Health Organization, 2020. Draft landscape of COVID-19 candidate vaccines – 9 September 2020. https://www.who.int/who-documents-detail/draft-landscape-of-COVID-19-candidate-vaccines. Accessed date: September 12, 2020

14. World Health Organisation, 2019. Ten Threats to Global Health in 2019. https://www.who.int/vietnam/news/feature-stories/detail/ten-threats-to-global-health-in-2019. Accessed date: September 12, 2020

15. Dubé E, Laberge C, Guay M, Bramadat P, Roy R, Bettinger J, 2013. Vaccine hesitancy: An overview. Human Vaccines and Immunotherapeutics. 9(8);1763–1773.

16. Ogundele OA, Ogundele T, Beloved O, 2020. Vaccine hesitancy in Nigeria: Contributing factors – way forward. The Nigerian Journal of General Practice, 18(1); 1–4.

17. Ghinai I, Willott C, Dadari I, Larson HJ, 2013. Listening to the rumours: What the northern Nigeria polio vaccine boycott can tell us ten years on. Global Public Health, 8(10);1138–1150.

18. Emmanuel Awucha N, Chinelo Janefrances O, Chima Meshach A, Chiamaka Henrietta J, Ibilolia Daniel A, Esther Chidiebere N, 2020. Impact of the COVID-19 Pandemic on Consumers’ Access to Essential Medicines in Nigeria. The American journal of tropical medicine and hygiene, 10.4269/ajtmh.20-0838.

19. Zhong BL, Luo W, Li HM, Zhang QQ, Liu XG, Li WT, Li Y, 2020. Knowledge, attitudes, and practices towards COVID-19 among Chinese residents during the rapid rise period of the COVID-19 outbreak: a quick online cross-sectional survey. Int J Bio Sci 16: 1745–1752.

20. Reluga TC, Bauch CT, Galvani AP, 2006. Evolving public perceptions and stability in vaccine uptake. Mathematical Biosciences. 204(2);185–198

21. Ophori EA, Tula MY, Azih AV, Okojie R, Ikpo PE, 2014. Current trends of immunization in Nigeria: prospect and challenges. Tropical medicine and health. 42(2);67–75.

22. Harmsen IA, Mollema L, Ruiter RA, Paulussen TG, de Melker H. E, Kok G. 2013. Why parents refuse childhood vaccination: a qualitative study using online focus groups. BMC public health, 13;1183.

23. Neumann-Böhme S, Varghese NE, Sabat I, Barros PP, Brouwer W, van Exel J, Schreyögg J, Stargardt T, 2020. Once we have it, will we use it? A European survey on willingness to be vaccinated against COVID-19. The European journal of health economics: HEPAC: health economics in prevention and care, 21(7);977–982.

24. IPSOS. 2020. Three in four adults globally say they would get a vaccine for COVID-19. https://www.ipsos.com/ipsos-mori/en-uk/three-four-adults-globally-say-they-would-get-vaccine-covid-19. Accessed date: September 12, 2020

25. Harapan H, Wagner AL, Yufika A, Winardi W, Anwar S, Gan AK, Setiawan AM, Rajamoorthy Y, Sofyan H, Mudatsir M. 2020. Acceptance of a COVID-19 Vaccine in Southeast Asia: A Cross-Sectional Study in Indonesia. Frontiers in public health, 8;381.

26. Wong LP, Alias H, Wong PF, Lee HY, AbuBakar S. 2020. The use of the health belief model to assess predictors of intent to receive the COVID-19 vaccine and willingness to pay. Human vaccines & immunotherapeutics. 1–11.

27. Gunnala R, Ogbuanu IU, Adegoke OJ, Scobie HM, Uba BV, Wannemuehler KA, Ruiz A, Elmousaad H, Ohuabunwo CJ, Mustafa M, Nguku P, Waziri NE, Vertefeuille JF. 2016. Routine Vaccination Coverage in Northern Nigeria: Results from 40 District-Level Cluster Surveys, 2014-2015. PloS one. 11(12);e0167835.

28. Oku A, Oyo-Ita A, Glenton C, Fretheim A, Eteng G, Ames H, Muloliwa A, Kaufman J, Hill S, Cliff J, Cartier Y, Bosch-Capblanch X, Rada G, Lewin S. 2017. Factors affecting the implementation of childhood vaccination communication strategies in Nigeria: a qualitative study. BMC public health, 17(1);200.

29. Ughasoro MD, Esangbedo DO, Tagbo BN, Mejeha IC. 2015. Acceptability and Willingness-to-Pay for a Hypothetical Ebola Virus Vaccine in Nigeria. PLoS neglected tropical diseases, 9(6);e0003838.

30. Chukwuocha UM, Okorie PC, Iwuoha GN, Ibe SN, Dozie IN, Nwoke BE. Awareness, perceptions and intent to comply with the prospective malaria vaccine in parts of South Eastern Nigeria. Malar J. 2018;17:187.

31. Marti M, de Cola M, MacDonald NE, Dumolard L, Duclos P. (2017). Assessments of global drivers of vaccine hesitancy in 2014-Looking beyond safety concerns. PloS one. 12(3);e0172310.

32. Adebisi YA, Eliseo-Lucero Prisno D III, Nuga B.B. 2020. Last fight of wild polio in Africa: Nigeria’s battle. Public Health in Practice, 1, 100043.

33. Thomson A, Watson M. 2012. Listen, understand, engage. Science translational medicine. 4(138);138ed6.

34. Adekunle AI, Adegboye OA, Gayawan E, McBryde ES. Is Nigeria really on top of COVID-19? Message from effective reproduction number. 2020. Epidemiol Infect. 148:e166.

35. Anderson RM, May RM. 1985. Vaccination and herd immunity to infectious diseases. Nature. 4;318(6044):323–9

